# Phase One Development of a Patient-Reported Outcome Measure for Low Anterior Resection Syndrome

**DOI:** 10.64898/2026.07.27.26359019

**Authors:** Alexandra Coxon-Meggy, Emily Farrow, Laura Knight, Ian Bissett, Liliana Bordeianou, Marylise Boutros, Jennifer Burch, Peter Christensen, Neil Corrigan, Julie Croft, Marie Demian, Claudia DiVenti, Sunny Dhadlie, Katrine J Emmertsen, Katie Gordon, Sarah Faris-Sabboobeh, Nicola Fearnhead, Julio Flavio Fiore, Celia Keane, Charles Knowles, Christina Lloydwin, Franco Marinello, Alun Meggy, Helen Mohan, Kheng-Seong Ng, Camila L. P. Oliveira, Lucia Oliveira, Aaron Quyn, Azmina Rose, Deborah Stocken, Andrea Warwick, Judith White, Julie Cornish

**Author notes:** Corresponding Author: Emily Farrow.

## Abstract

**Background:** The Low Anterior Resection Syndrome (LARS) score is an internationally validated instrument for identifying bowel dysfunction following anterior resection for rectal cancer. Although widely used, it has been shown to have limited sensitivity for capturing the impact of LARS on daily-life and response to treatment. We have therefore developed a novel patient-reported outcome measure (PROM): the LARS Impact and Consequences Assessment Tool (LARS-ICAT).

**Methods:** Initial development of LARS-ICAT followed a five-stage process following established PROM development guidance. Stage one established the conceptual foundation through previously published Delphi consensus. Stage two involved item generation, followed by evaluation of content validity through patient focus groups (n=11) in stage three. Stage four comprised iterative expert review and refinement through clinical consensus with patient involvement. Stage five involved cognitive interviews with patients conducted across five rounds (n=23).

**Results:** Several items identified through the Delphi consensus were reworded as they included multiple concepts. A one-month recall period was selected, with six and four response options for symptom and consequence items respectively. Additional consequence items, including impact on sleep and transport use, were incorporated. Focus groups and clinicians emphasised the importance of capturing individual symptom burden, leading to the addition of symptom bother scales. These iterative refinements culminated in LARS-ICAT v2.6.

**Conclusions:** LARS-ICAT is a novel PROM designed to assess symptom burden and treatment response in LARS. Future studies will assess its psychometric properties. Once validated, LARS-ICAT will provide a comprehensive, patient-centred assessment of LARS, enhancing our ability to manage this challenging condition.

**Strengths and Limitations:**

- The LARS-ICAT was specifically designed to measure change over time and responsiveness to treatment, addressing important limitations of existing instruments, such as the LARS score.
- Input from patients and experts across multiple countries improved the content validity and cross-cultural relevance of the PROM.
- This work also led to the establishment of the LARS Collaborative, an international research group dedicated to advancing education, engagement and understanding of LARS and its management.
- The study included only English-speaking participants, which may limit the generalisability of the instrument to non-English-speaking populations until formal translation, linguistic validation and cross-cultural validation has been undertaken.

## Introduction

Low anterior resection syndrome (LARS) encompasses a spectrum of debilitating bowel symptoms that commonly arise after rectal resection (1). These symptoms affect up to three-quarters of patients within the first-year post-surgery and persist in approximately 40% of patients over the longer term (2). Consequently, LARS has become a major survivorship issue following rectal cancer surgery. This burden is amplified by improving survival and the rising incidence of rectal cancer in younger populations (3,4)(5), rendering the impact of bowel dysfunction on quality of life (QoL) a critical clinical concern.

Two validated patient-reported outcome measures (PROMs) currently exist for LARS: the Memorial Sloan Kettering Colorectal Clinic Bowel Function Index (MSKCC-BFI) and the LARS Score (6)(7). While these instruments demonstrate strong concordance, the MSKCC-BFI has limited uptake, whereas the LARS Score has been widely adopted, translated and validated in over 20 languages (8)(9)(10)(11). However, the LARS Score was designed primarily as a screening instrument and lacks sensitivity to longitudinal change and treatment response, limiting its suitability for interventional trials (7)(12).

In 2020, an international Delphi consensus involving LARS experts and patients refined the definition of LARS, highlighting its substantial impact on quality of life and the importance of capturing individual experiences (13). The resulting definition encompassed eight symptoms and eight consequences, reflecting both functional impairment and broader psychosocial impact. Both the MSKCC-BFI and LARS Score predate the 2020 consensus definition and therefore exhibit incomplete content validity relative to the current conceptualisation of LARS, particularly regarding obstructive symptoms and psychosocial consequences (2).

These limitations highlight the need for a new PROM that reflects contemporary understanding of LARS and that demonstrates sensitivity to change. In addition to providing a robust outcome measure for research, such a tool could support tailored clinical management by helping to identify the symptoms and consequences most important to individual patients. This paper reports the initial phase of development of the LARS Impact and Consequences Assessment Tool (LARS-ICAT), a new PROM designed for use across English-speaking populations. Specifically, it focuses on item generation and content validity evaluation to establish a robust foundation for its use in both clinical practice and research.

## Methodology

The development of the LARS-ICAT followed established guidelines for PROM development, including the International Society for Pharmacoeconomics and Outcomes Research (ISPOR) good research practices and FDA guidance on content validity (14)(15)(16,17). In addition, the Consensus-based Standards for the selection of health measurement instruments (COSMIN) framework was used to guide evaluation of content validity during Stages 3 and 5 (18)The initial development phase consisted of five sequential stages, summarised below (Figure 1). Stage one is not described in this publication, as it has been reported previously (19).

**Figure 1.**
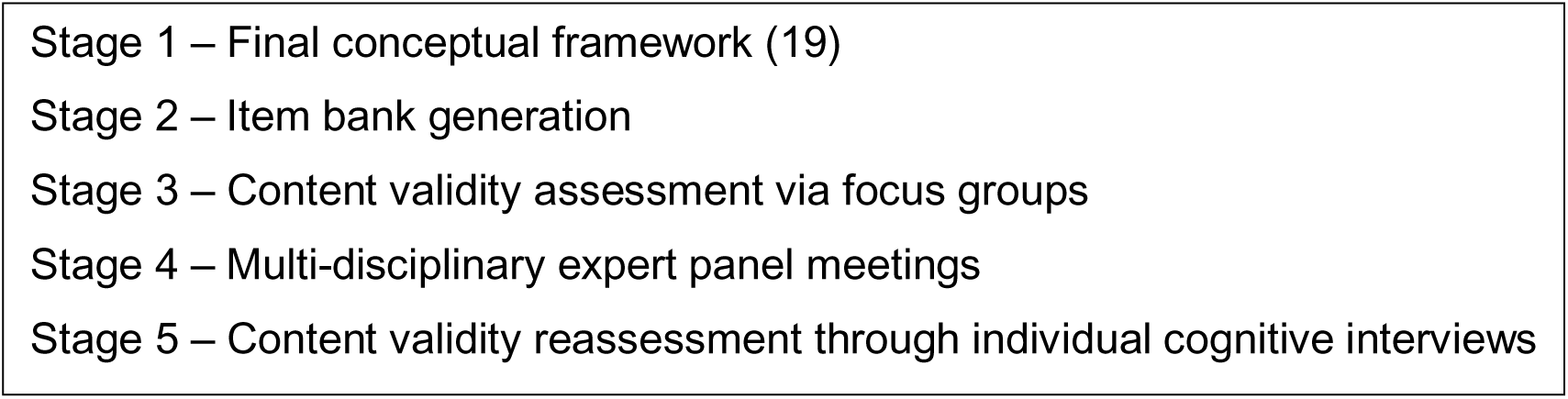
The initial development phase of LARS-ICAT PROM outlined in five sequential stages.

### Study Management Group and Patient Involvement

The Study Management Group (SMG) provided methodological and clinical oversight throughout LARS-ICAT development. It comprised international LARS experts, including the study group for Pathway of Low Anterior Resection Syndrome Relief after Surgery (POLARiS) and contributors to the 2020 Consensus Definition and the LARS score, alongside qualitative methodologists and patient representatives. Governance subsequently evolved into the LARS Collaborative, an international, multidisciplinary research consortium advancing global LARS research and clinical practice.

### Inclusion and exclusion criteria

Eligible patients for Stage 3 and 5 of the study were adults (>18 years) who had undergone anterior resection for rectal cancer and had restored bowel continuity at least 6 months previously. As the LARS-ICAT was developed in English, participants were required to be sufficiently proficient in spoken and written English. Exclusion criteria included concurrent adjuvant therapy, recurrent or metastatic disease, physical or cognitive limiting meaningful participation.

### Stage 2: Item Generation

The SMG undertook item generation for the LARS-ICAT, using the eight symptoms and eight consequences identified through the international Delphi consensus on LARS as the underlying conceptual framework (Stage 1). Items and response options were developed to reflect these domains, with additional content generated and refined through an iterative consensus process with the SMG. This approach ensured comprehensive representation of the multidimensional impact of LARS while maintaining fidelity to the consensus-derived construct.

### Stage 3: Focus Groups

Focus groups were conducted in selected English-speaking countries to assess content validity in accordance with COSMIN guidance, using a topic guide structured around the COSMIN criteria for relevance, comprehensibility and understandability (20). Participants were identified through institutional clinical databases and previous research who had consented to future contact. Eligible individuals were invited via post or email.

Three online focus groups were held: two in the UK (n=5, n=3) and one with participants from Australia and North America (n=3). Participants received the preliminary LARS-ICAT in advance. Consistent with COSMIN methodology, sample size was determined by the need to generate sufficiently rich data and achieve adequate coverage of key content validity domains rather than a predefined target (18). Focus groups were facilitated by trained qualitative researchers and clinicians (LK, CK, ACM, CL), recorded, transcribed verbatim and analysed by two researchers using a question-feature coding, pattern-coding and cross-group comparison. Findings were reviewed iteratively by the SMG to inform instrument refinement, with a consolidated summary circulated for consensus.

### Stage 4: Multi-disciplinary Meetings

A series of multi-disciplinary international meetings, conducted both face-to-face and virtually between December 2023 and September 2025, supported the iterative refinement of LARS-ICAT (Figure 2). Attendees included international clinicians, researchers and PROM methodologists and patient representatives from the SMG (Figure 3 & 4). Both focus group and cognitive interview findings were systematically reviewed, and proposed amendments to items, response options and formatting were discussed. Changes to LARS-ICAT were implemented following formal consensus among the panel, ensuring that revisions were clinically relevant and aligned with patient priorities.

**Figure 2.**
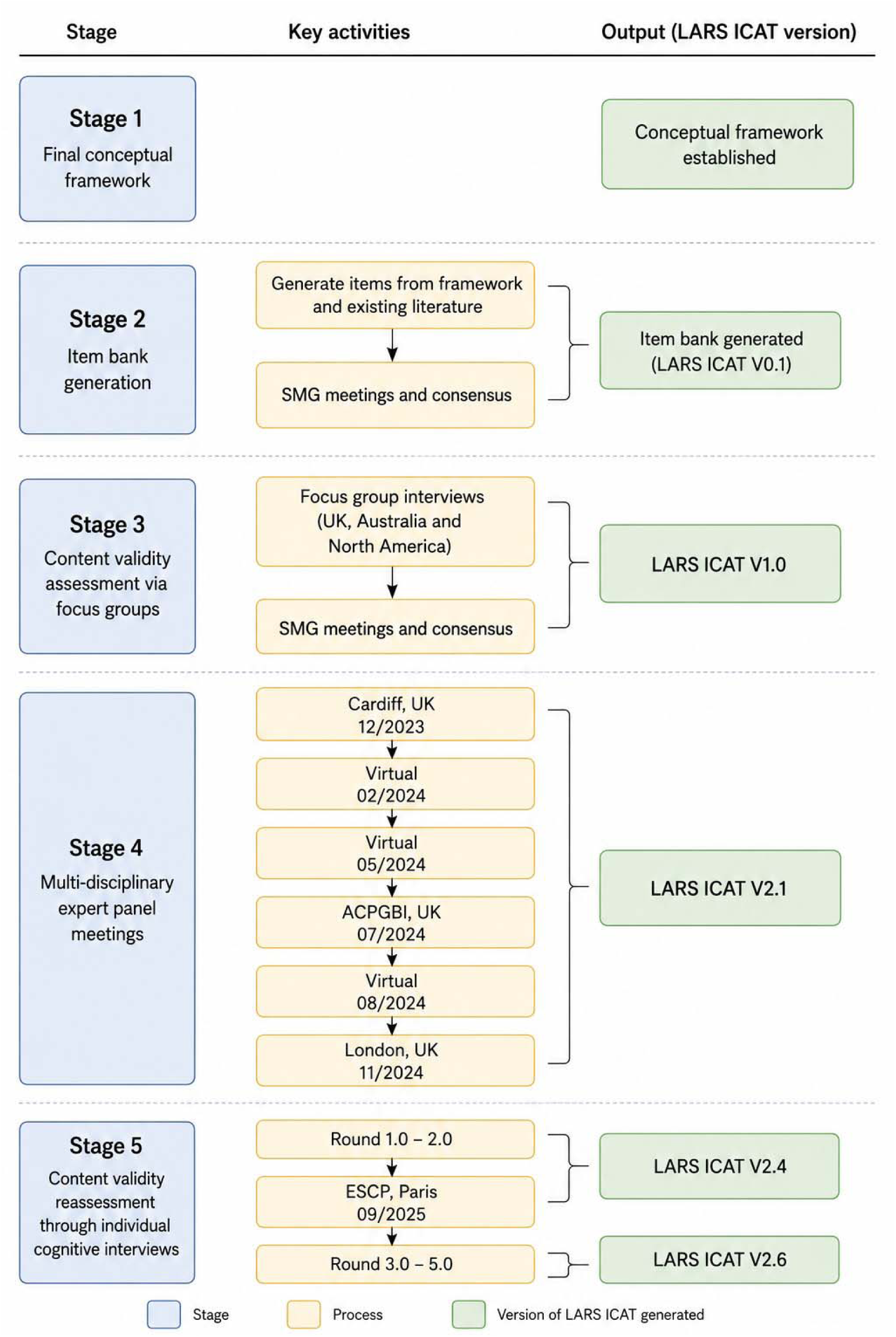
Overview of Phase 1 LARS-ICAT development and content validation, showing the five study stages and corresponding instrument versions.

**Figure 3.**
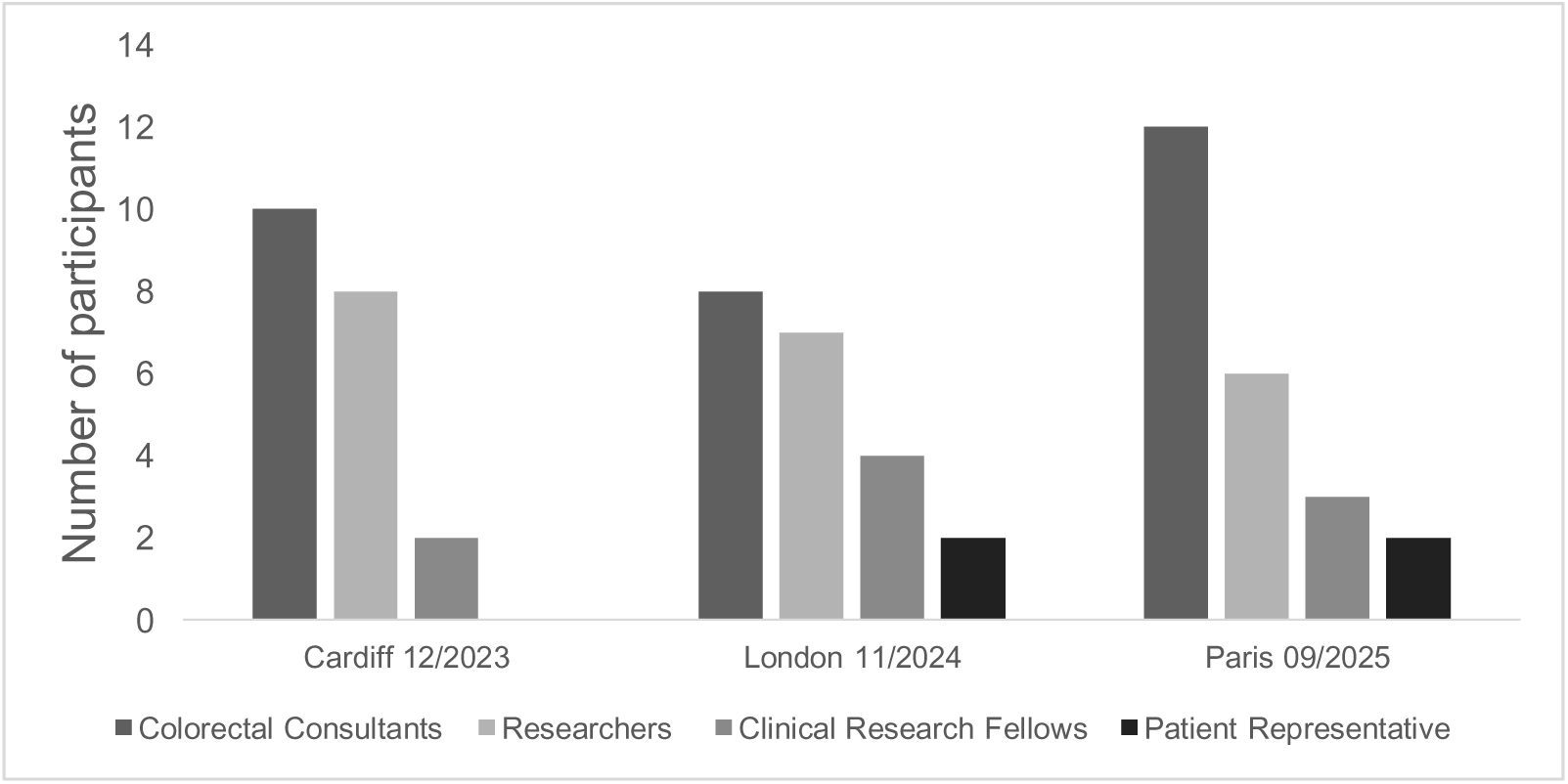
The multidisciplinary expertise represented across Stage Three face-to-face meetings.

**Figure 4.**
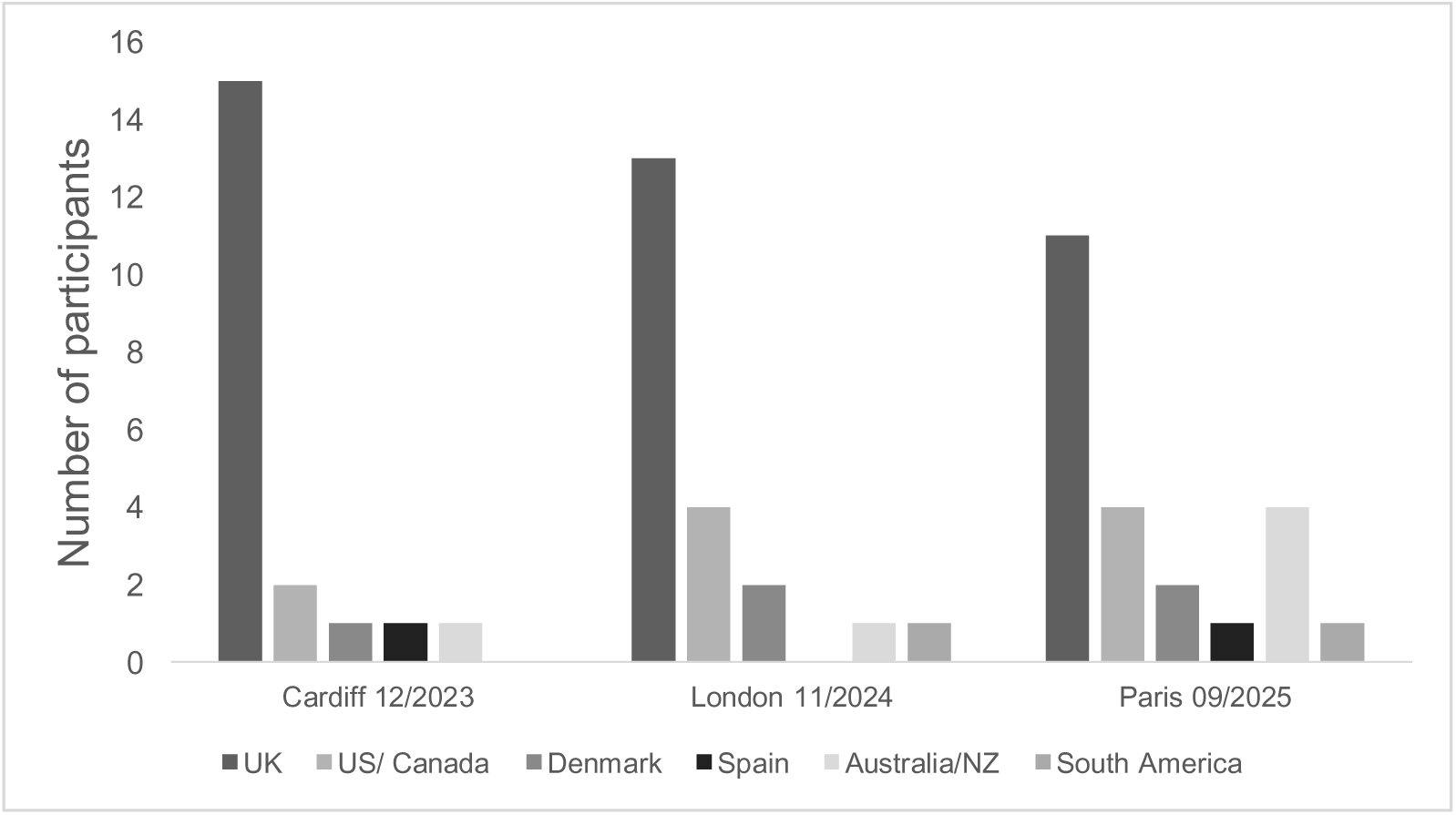
The international representation across Stage Three face-to-face meetings.

### Stage 5: Cognitive Interviews

Cognitive interviews were conducted in selected English-speaking countries to reassess the comprehensibility and relevance of the LARS-ICAT. UK participants were recruited through a LARS support group, while in New Zealand, Australia and the United States patients were recruited through follow-up clinics.

A semi-structured interview guide was pilot tested with two patients. Interviews were conducted by experienced qualitative researchers or clinicians (LK, CK, EF, SD, CD, CO) and audio recorded. Interviews were conducted iteratively in multiple rounds and findings reviewed by the SMG. Interviews continued until thematic saturation was achieved, ensuring comprehensive coverage of patient experiences and no emergence of new insights that would lead to further revisions of the instrument.

### Data Collection

Demographic and clinical data, including each participant’s LARS score, were collected to contextualise symptom burden. Focus groups and cognitive interviews were conducted via Microsoft Teams, with video and audio recordings taken respectively and transcribed verbatim. Transcripts were securely stored in line with the local sponsor’s data management policies, ensuring compliance with institutional governance and data protection standards.

### Ethical approval

Stage 3 was approved by Wales Research and Ethics Committee (REC) on 11^th^ July 2022 (reference 22/WA/0213) and registered on public data base (clinicaltrials.gov; NCT05605600). Stage 5 received ethical approval from Cardiff University, School of Medicine REC on 4^th^ March 2025 (SREC reference 25/01), Austin Health Human REC, Australia on 23^rd^ March 2026 (HREC/28699/Austin-2025), the Cleveland Clinic Institutional Review Board (24-1093) and Auckland Health REC on 18^th^ November 2025 (AH24533).

## Results

### Stage 2: Item Generation

Using the eight symptoms and eight consequences identified through the international Delphi consensus as the conceptual framework (21), the SMG generated an initial 18-item draft instrument (LARS-ICAT v1.0). To enhance its clinical utility and ensure comprehensive assessment, additional items were included to capture usual stool type (using Bristol Stool Chart) and overall QoL impact. Symptom items employed a seven-point frequency-based response scale, while consequence items used a four-point impact scale anchored by visual face symbols. No consensus was reached regarding the optimal recall period at this stage. The resulting draft instrument provided the basis for subsequent content validity evaluation through focus groups.

### Stage 3: Focus Groups

Participant characteristics are presented in **Error! Reference source not found.**. Issues and observations identified during focus groups were systematically mapped against the ten COSMIN criteria for content validity, assessing relevance, comprehensiveness and understanding (Table 2).

**Table 1.** Participant characteristics by focus group.

| **Characteristic** | **FG1 (n = 5)** | **FG2 (n = 3)** | **FG3 (n = 3)** |
| --- | --- | --- | --- |
| Sex (male:female) | 2:3 | 1:2 | 1:2 |
| Age (years) |  |  |  |
| 51-60 | 4 | 1 | 1 |
| 61-70 | 1 | 1 | 2 |
| 71-80 | 0 | 1 | 0 |
| Employment |  |  |  |
| Full time | 3 | 1 | 0 |
| Part time | 0 | 1 | 0 |
| Retired | 2 | 1 | 3 |
| Years since surgery |  |  |  |
| 0-5 | 1 | 3 | 0 |
| 5-10 | 3 | 0 | 3 |
| >10 | 1 | 0 | 0 |
| Ileostomy | 5 | 1 | 2 |
| Radiotherapy | 0 | 0 | 1 |
| Chemotherapy | 2 | 0 | 1 |
| LARS Score |  |  |  |
| No | 0 | 1 | 1 |
| Minor | 1 | 2 | 0 |
| Major | 4 | 0 | 2 |

*FG = focus group, LARS = Low Anterior Resection Syndrome*

**Table 2.**
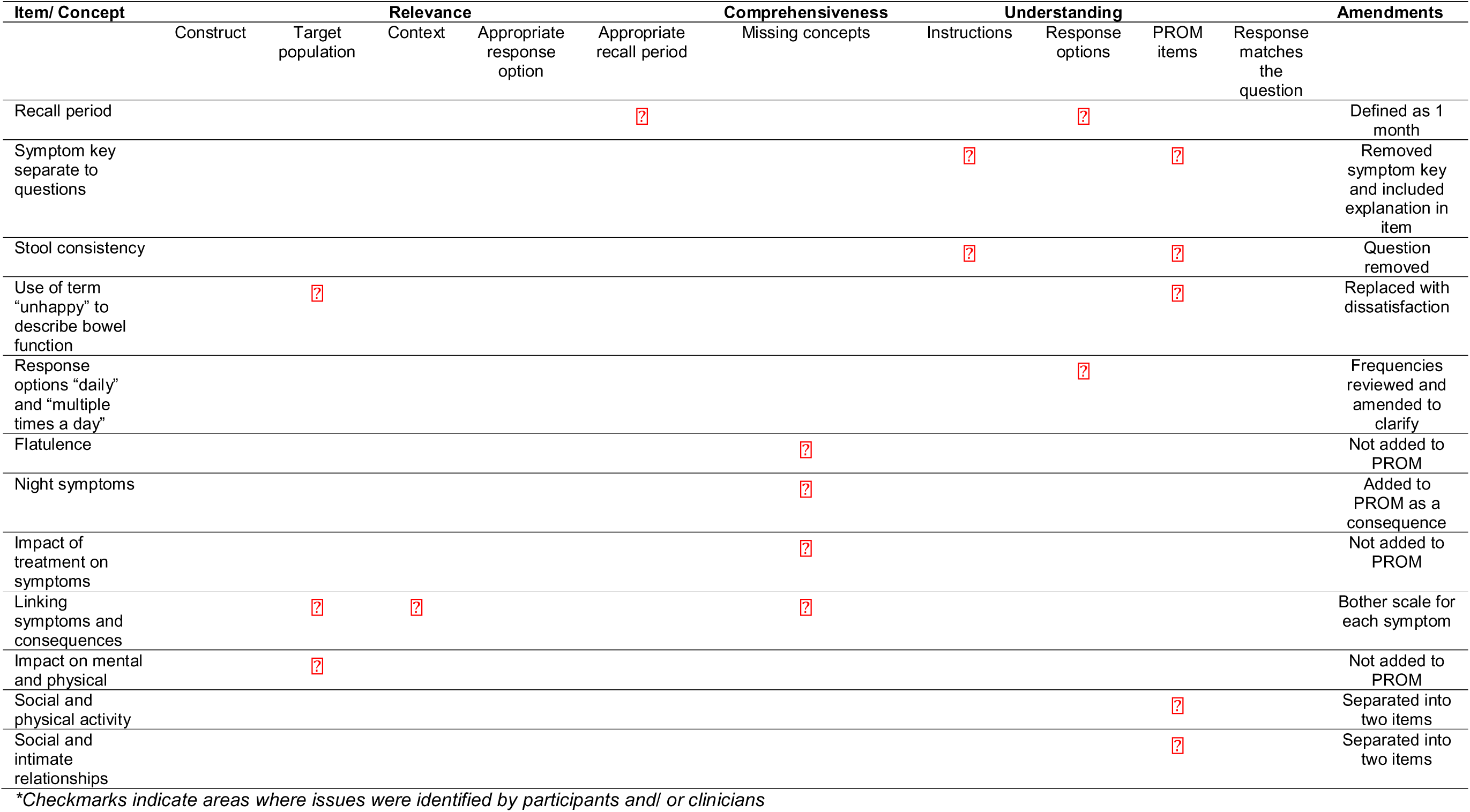
Summary of focus group findings mapped to COSMIN content validity domains and resulting amendments to the LARS-ICAT.

Achieving consensus for the recall period was challenging. Participants noted high variability in bowel habits, making longer recall periods difficult without a bowel diary:

> *‘I myself would not remember over a month; it’s such a day-to-day thing, I could tell you how I was last week, but a month, I’d have to track.’ (NLS010, focus group 3)*

Participants highlighted that the absence of a defined recall period could bias responses towards symptomatic fluctuations. Frequency response options were also challenged, particularly where categories were inconsistent with item content such as “increased stool frequency” “multiple times a day”. These findings highlighted the need for careful consideration of the recall period and response scaling.

Flatulence and nocturnal symptoms were identified as missing from LARS-ICAT v1.0. SMG discussions focused on the optimal classification of nocturnal symptoms, considering whether they should be represented as a distinct symptom or consequence item or applied as a modifier to all symptom items. Although flatulence was also identified as a missing concept, it was not incorporated into the LARS-ICAT, consistent with its exclusion from the Delphi consensus. As the Delphi process was adequately powered, its findings were considered more definitive.

A recurring theme was the importance of capturing the broader impact of symptoms. Many participants expressed that their lived experiences had often been overlooked or inadequately addressed in clinical consultations. They suggested that explicitly linking specific symptoms to their psychosocial consequences would enable more meaningful communication with clinicians:

> *‘It would make your conversation with the clinician much more meaningful if you could explain that it’s this one symptom that’s causing all these consequences and really having an impact on you.’* (NLS002, focus group 1)

Following qualitative analysis of the focus group transcripts, a comprehensive summary of findings was produced and presented at international multi-disciplinary meetings, where implications of findings were discussed.

### Stage 4: Multi-disciplinary Consensus Meetings

Focus group findings were discussed at consensus meetings and informed item retention and modification, resulting in LARS-ICAT v1.6 (Tables 3 & 4). A one-month recall period was agreed to capture typical symptom patterns while minimising transient symptomatic fluctuations. Frequency response options were revised to a six-point scale, reflecting clinician preference for granularity without a neutral midpoint.

Focus group participants and clinicians highlighted the importance of linking symptoms with consequences. An 11-point severity-bother scale was therefore introduced for each symptom (LARS-ICAT v1.8) to capture the degree of bother caused by individual symptoms. Although some clinicians expressed concerns about response burden, analytical complexity and cross-cultural interpretation of “bother”, the scale was retained for psychometric evaluation.

Additional items were added following focus group and clinical consensus with patient involvement to strengthen content validity. Consequence items addressing sleep and public transport were added, while “most common stool consistency” was removed because of ambiguity (Table 3 & 4). The original approach of applying “Do you have this symptom at night?” as a modifier to each symptom item was revised and consolidated into a single consequence item to reduce response burden and avoid redundancy.

**Table 3.**
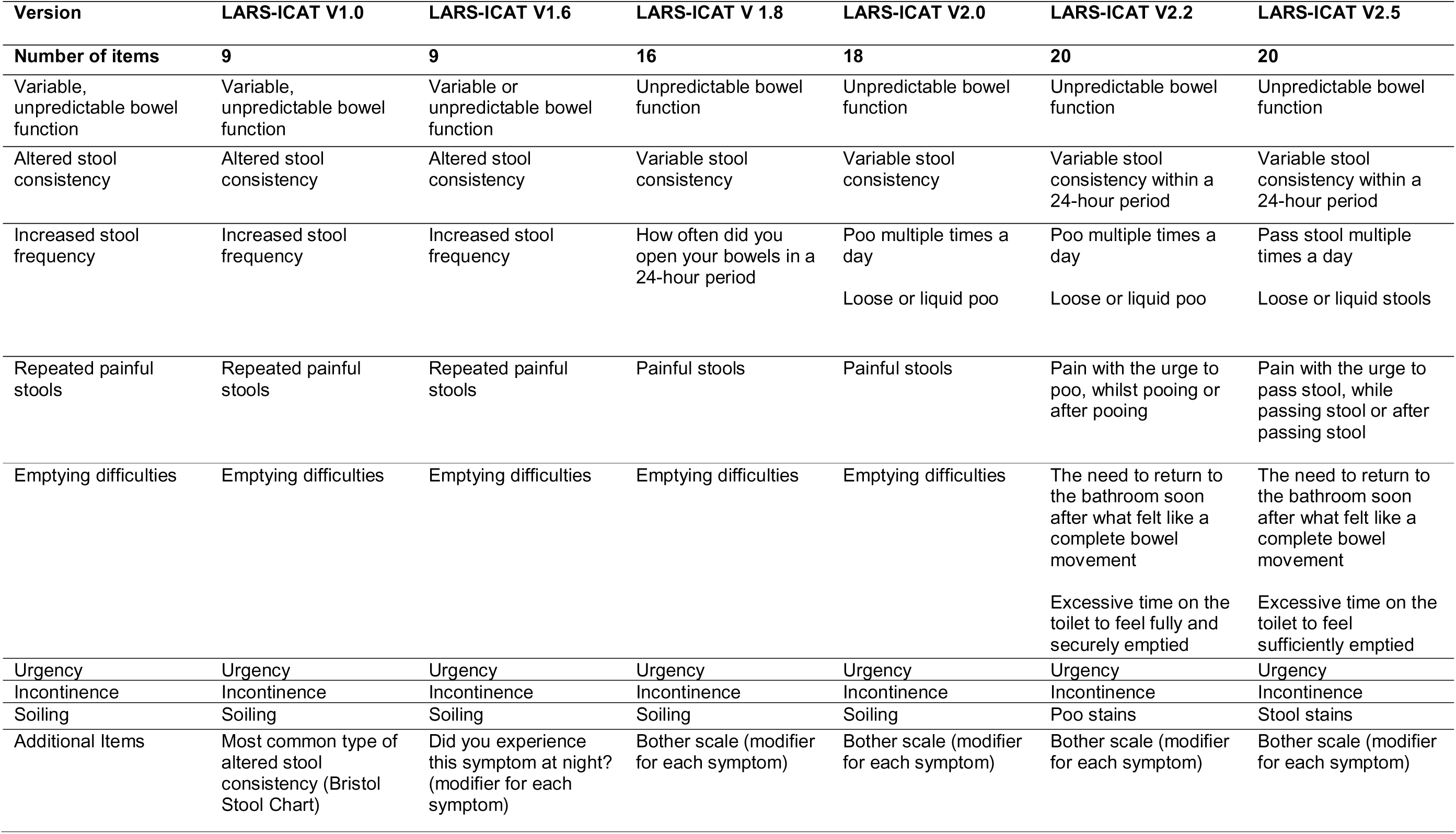
Iterative rewording of symptom items in LARS-ICAT.

Methodologists recommended refining items to measure a single concept. Consequently, items such as “relationships and intimacy” and “social and physical activities”, were revised or separated to improve clarity and measurement precision (Table 4). These iterative refinements resulted in LARS-ICAT v2.1 comprising 30 items for evaluation in cognitive interviews.

**Table 4.**
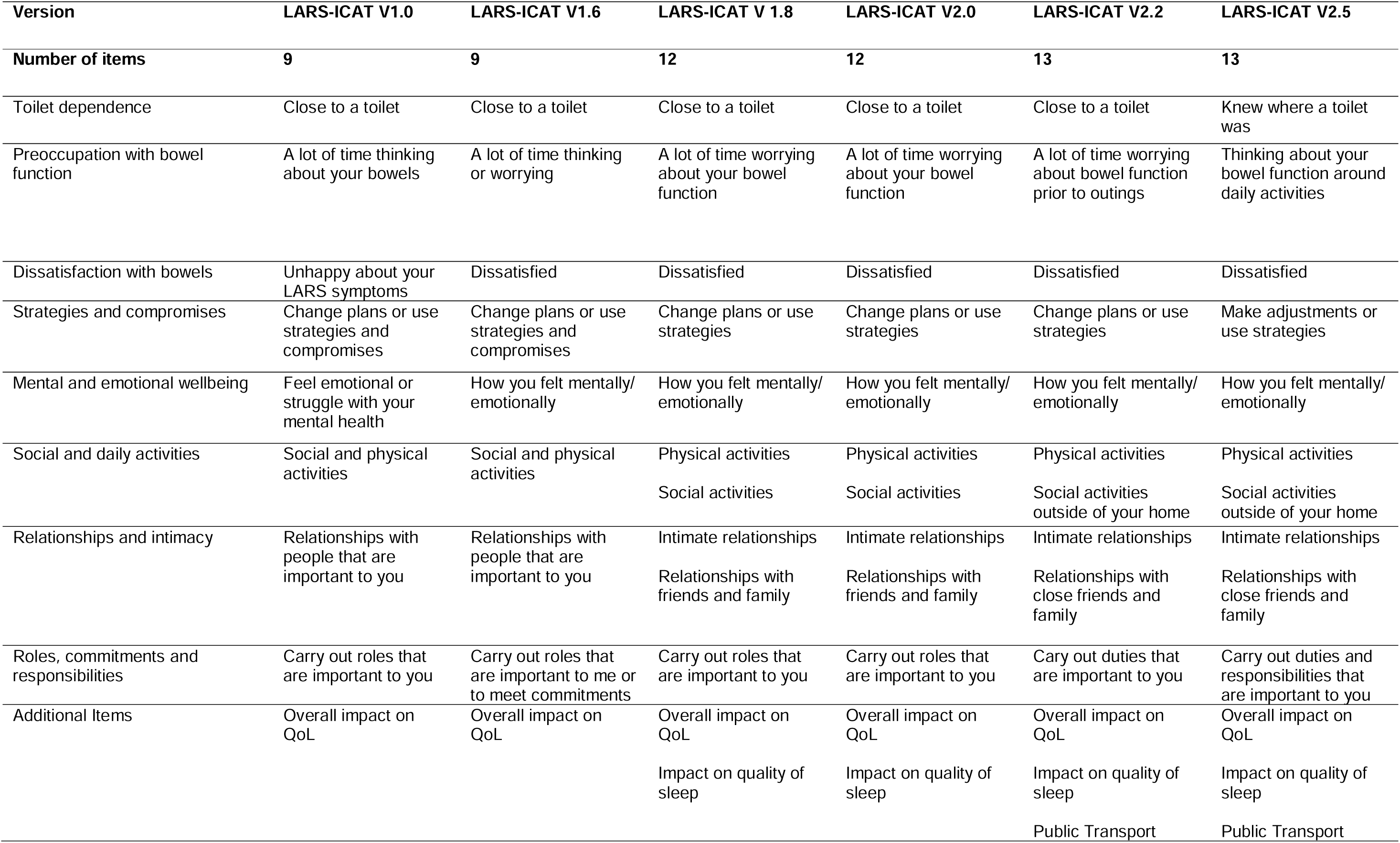
Iterative rewording of consequence items in LARS-ICAT.

### Stage 5: Cognitive Interviews

Twenty-three international cognitive interviews were conducted over five rounds in the UK, US, New Zealand and Australia. The one-month recall period was generally acceptable, although some participants reported difficulty in recalling symptoms over this timeframe. In Round 1 participants frequently overlooked the “last month” instruction, prompting formatting changes to emphasise the recall period. More bothersome symptoms, such as faecal incontinence, and consequence items were more readily recalled over one month.

Iterative rewording improved clarity (Table 3 & 4). The distinction between “soiling” and “incontinence” was confusing; “soiling” was replaced with “staining”, which was better understood. “Variable stool consistency” was clarified as “within 24 hours”, enabling consistent responses. Participants in the US reported the term “poo” was perceived as infantilising. Consequently, this terminology was replaced throughout the instrument with “stool” or “bowel movement”.

Participants in the UK and Australia consistently interpreted the bother scale as reflecting the impact of symptoms on daily life, including interference and emotional distress, and were comfortable using the full 0-10 Likert scale. In contrast, participants in the US and New Zealand found the term “bother” less intuitive.

Nevertheless, the concept of “bother” is widely used in existing PROMS (22)(23)(24). Although concerns regarding the use of the bother scale were raised at Stages 4 and 5, psychometric evaluation will provide an objective assessment of its performance.

Participants in the UK, New Zealand and Australia found the item “Did you feel dissatisfied with your bowel function?” ambiguous. In LARS-ICAT v2.1, its placement immediately before the QoL impact question led participants to perceive both as global assessments, prompting its relocation among other consequence items. However, participants continued to find the concept of bowel function “satisfaction” difficult to interpret, with many noting they would not have described their bowel function in these terms even before surgery. As this consequence was identified in the original Delphi process, the item was retained pending psychometric evaluation.

Following five rounds of international cognitive interviews LARS-ICAT v2.6 was generated.

**Table 5.** Participant characteristic by cognitive interview round.

|  | Round 1<br>(n=6) | Round 2<br>(n=3) | Round 3<br>(n=4) | Round 4<br>(n=8) | Round 5<br>(n=2) |
| --- | --- | --- | --- | --- | --- |
| LARS-ICAT Version | 2.1 | 2.2 | 2.4 | 2.5 | 2.6 |
| Sex (M:F) | 3:3 | 2:1 | 3:1 | 1:7 | 0:2 |
| Age, years; mean<br>(range) | 64 (53-73) | 65 (57-72) | 66 (54-75) | 54 (30-76) | 58 (53-63) |
| Nationality | UK (6) | UK (3) | US (4) | US (3)<br>New Zealand (3)<br>Australia (2) | UK (2) |
| Years since bowel<br>continuity |  |  |  |  |  |
| < 2 | 0 | 0 | 2 | 3 | 0 |
| 2-5 | 1 | 2 | 2 | 3 | 1 |
| 5-10 | 5 | 1 | 0 | 2 | 1 |
| Ileostomy | 4 | 3 | 0 | 6 | 1 |
| Radiotherapy | 2 | 1 | 0 | 3 | 0 |
| Chemotherapy | 3 | 1 | 0 | 3 | 1 |
| LARS Score |  |  |  |  |  |
| No | 0 | 1 | 0 | 0 | 0 |
| Minor | 1 | 0 | 2 | 3 | 0 |
| Major | 5 | 2 | 2 | 5 | 2 |

## Discussion

The LARS-ICAT is a novel PROM developed to assess the symptoms and consequences of LARS following rectal resection. This study utilised qualitative research methods to develop a PROM based on the refined definition of LARS (13). Maintaining a high fidelity to the Delphi Consensus work was a priority. Assessment of content validity was performed through focus groups and cognitive interviews with a total of 34 participants from the UK, USA, Australia and New Zealand.

Care was taken to ensure alignment with COSMIN criteria and FDA guidance for content validity (25)(26). The LARS-ICAT was developed with extensive patient involvement from multiple English-speaking countries, with the use of focus groups and cognitive interviews to capture the lived experience of LARS. Previously missing concepts, such as nocturnal symptoms, travel and symptom-related bother were identified and incorporated into the instrument. Input from patients and experts across countries improved the generalisability and cross-cultural relevance of the PROM. This work also led to the establishment of the LARS Collaborative, an international research group advancing education, engagement and understanding of LARS and its management.

Despite the methodological rigour applied in developing LARS-ICAT, several limitations should be acknowledged. First, the study included only English-speaking participants, which may limit the generalisability of the instrument to non-English-speaking populations until formal linguistic and cultural validation is completed. A protocol outlining a robust process for translation and validation is in development and will be implemented after the next stage of validation to incorporate multiple language options. Second, the small sample sizes of focus groups and cognitive interviews may not capture the full heterogeneity of the LARS experience, particularly concerning ethnicity and educational background. Finally, coordinating local ethical approvals and conducting cognitive interviews across multiple continents proved logistically challenging. As a result, the cognitive interview groups were not equal in size or geographical distribution. Nevertheless, the data generated from these interviews were qualitatively rich and provided substantial insight, contributing significant value to the development of the LARS-ICAT.

The LARS-ICAT addresses key limitations of prior instruments, such as the LARS score. Unlike the LARS score, which was primarily developed as a screening tool, the LARS-ICAT is specifically designed to assess longitudinal change and treatment response. Importantly, the LARS-ICAT aligns with the revised 2020 consensus definition of LARS, providing a more comprehensive and contemporary measure of the condition. From a research perspective, the LARS-ICAT provides a validated framework for assessing symptom burden and functional impact in longitudinal studies and clinical trials. The LARS-ICAT also enables more consistent evaluation of interventions for LARS, supporting meta-analyses and cross-study comparisons, ultimately advancing both clinical and research efforts in this area.

The LARS-ICAT now requires formal psychometric validation to evaluate its measurement properties, including reliability, construct validity and responsiveness. This validation is currently being undertaken as part of the Psychometric Evaluation of LARS-ICAT (PELICAT study IRAS 314213), which will recruit English-speaking participants from multiple international sites to ensure robust, cross-cultural assessment of the instrument. Regulatory approval will be sought be each contributing institution and data will be combined for analysis and prepared as a separate manuscript.

## Data Availability

All data produced in the present study are available upon reasonable request to the authors

## Patient and Public Involvement

Patient representatives with lived experience of LARS contributed to the development of the LARS-ICAT through participation in multidisciplinary meetings. They reviewed the preliminary instrument, advised on the relevance and comprehensiveness of its content and highlighted concepts that had been under-represented. Their contributions informed the inclusion and refinement of items, thereby strengthening the instrument’s content validity. In addition, patients from multiple English-speaking countries participated in focus groups and cognitive interviews to assess the relevance, clarity and acceptability of the instrument. A patient representative was involved in the dissemination of the study findings and is a co-author of this manuscript.

